# Genome-wide association study of REM-sleep behaviour disorder identifies new risk loci

**DOI:** 10.64898/2025.12.01.25339623

**Authors:** Emma N. Somerville, Lang Liu, Lynne Krohn, Farnaz Asayesh, Jamil Ahmad, Dan Spiegelman, Meron Teferra, Manuela Tan, Sheena Waters, Cristina Simonet, Laura Pérez-Carbonell, Anette Schrag, Pauline Dodet, Isabelle Arnulf, Michele T.M. Hu, Yo-El Ju, Jacques Y. Montplaisir, Jean-François Gagnon, Alex Desautels, Yves Dauvilliers, Merve Aktan-Süzgün, Abubaker Ibrahim, Ambra Stefani, Birgit Hogl, Carles Gaig, Angelica Montini, Gerard Maya, Alex Iranzo, Monica Serradell, Gian Luigi Gigli, Mariarosaria Valente, Francesco Janes, Andrea Bernardini, Karel Sonka, David Kemlink, Petr Dusek, Gültekin Tamgüney, Michael Sommerauer, Michela Figorilli, Monica Puligheddu, Brit Mollenhauer, Claudia Trenkwalder, Friederike Sixel-Doring, Giuseppe Plazzi, Francesco Biscarini, Elena Antelmi, Valerie Cochen De Cock, Wolfgang Oertel, Annette Janzen, Michele Terzaghi, Giuseppe Fiamingo, Anna Heidbreder, Christelle Charley Monaca, Luigi Ferini-Strambi, Kristina Kulsarova, Miriam Ostrozovicova, Matej Skorvanek, Femke Dijkstra, Mineke Viaene, Jitka Bušková, Beatriz Abril, Beatrice Orso, Pietro Mattioli, Dario Arnaldi, Bradley F. Boeve, Guy A. Rouleau, Ronald B. Postuma, Jaeyoon Chung, Daria Prilutsky, Kajsa Atterling Brolin, Alastair J. Noyce, Owen A. Ross, Cornelis Blauwendraat, Ziv Gan-Or, the Global Parkinson’s Genetics Program (GP2) the NAPS Consortium

## Abstract

Rapid-eye-movement (REM)-sleep behaviour disorder (RBD) can be a prodromal stage of α-synucleinopathies, including Parkinson’s disease (PD) and dementia with Lewy bodies (DLB), and provides a unique opportunity to study and understand early neurodegeneration. Previous research on RBD has identified common risk loci in several PD and DLB genes, but the full extent of its genetic architecture and disease-specific associations remain unknown.

In the present study, we aimed to identify novel loci associated with RBD by conducting an updated European RBD genome-wide association study (GWAS). We performed a meta-analysis using a previous RBD GWAS and newly genotyped RBD patients and controls, resulting in a total of 3,564 RBD cases and 140,393 controls analyzed. We further assessed the functional impact of new associations using fine-mapping and colocalization analyses with adult brain expression quantitative trait loci (eQTLs). To confirm if novel RBD loci are implicated in additional synucleinopathy phenotypes, we examined common variant associations in GWASs of PD risk, DLB risk, PD with RBD risk, PD age at onset, and various measures of progression. Additionally, rare variants were analyzed with the optimal sequence kernel association test (SKAT-O) in PD and DLB whole genome sequencing cohorts. Lastly, we analyzed known PD- and DLB-risk loci for associations with RBD.

We identified *RCOR1* as a novel risk locus for RBD, with fine-mapping suggesting the association to be driven by a variant in intron 2. Colocalization revealed no evidence for shared genetic signals with brain eQTLs, although this may reflect a lack of statistical power to detect an association. Common and rare variants in the *RCOR1* locus were not associated with PD, DLB, and additional α-synucleinopathy phenotypes. We observed several PD and DLB risk loci to be associated with RBD, including *MAPT*, *STK39*, and *SIPA1L2*. The *MAPT* associated variant tags the protective H2 haplotype, consistent with previous findings in PD. Several *SNCA* variants previously associated with PD or DLB were also associated with RBD, but with differing directions of effect, highlighting the complex genetic landscape underlying α-synucleinopathy subtypes. This study identifies new RBD loci and strengthens our understanding of the genetic modifiers underlying RBD. Further replication and functional studies will be required to validate our findings and explore their biological implications.

## Introduction

Treatment of *α*-synucleinopathies, including Parkinson’s diseases (PD), dementia with Lewy bodies (DLB), and multiple system atrophy (MSA), is marked by a clear lack of therapeutic options addressing disease progression. While current treatments have some success in alleviating symptoms, there is still a notable lack when it comes to interventions that can slow or stop the course of neurodegeneration for these disorders. An important reason for this gap is that by the time symptoms have become pronounced enough for an overt *α*-synucleinopathy diagnosis to be made, neurodegeneration has already progressed substantially.^1^

However, a subset of patients with *α*-synucleinopathies share a prodromal condition, isolated/idiopathic REM-sleep behaviour disorder (iRBD), that may be an answer to this problem. iRBD is a parasomnia characterized by dream enactment behaviour and loss of muscle atonia. iRBD is now recognized as a prodromal α-synucleinopathy, with more than 80% of diagnosed individuals eventually phenoconverting within 10-15 years of onset.^2–4^ Roughly 45-50% of subjects phenoconvert to PD, 45-50% phenoconvert to DLB, and 5% phenoconvert to MSA.^5–7^ Thus, iRBD is an early manifestation of these disorders, and provides an important opportunity to understand the mechanisms underlying neurodegeneration at its earliest stages. About 1-2% of the general adult population has iRBD, yet most of them are undiagnosed.^8–10^

Genetic research of iRBD is still in its early stages. Candidate gene analyses have been the predominant method utilized thus far^11–16^, and recently, the first RBD genome-wide association study (GWAS) in Europeans identified associations with *SNCA*, *GBA1*, *TMEM175*, *INPP5F*, and *SCARB2*.^17^ Although this study successfully provided insight into common genetic variation underlying RBD on a genome-wide scale, there is still a lack of power to detect associated variants of rarer allele frequencies and smaller effect sizes. To this end, we performed an updated RBD GWAS including the summary statistics from the original study and an additional 721 iRBD patients and 761 controls.

## Materials and methods

### Study population

The study population consisted of two cohorts. The first is the entire meta-analysis cohort from the previous RBD GWAS^17^, consisting of 2,843 iRBD and PD with probable RBD (PD+pRBD) cases and 139,636 controls. Briefly, 1,061 iRBD cases and 871 controls from this previous analysis were recruited by the International RBD Study Group (IRBDSG) from multiple centers across Europe and North America, while 7,515 controls were included from external cohorts including the HYPERGENES Project^18^, the Wellcome Trust Case Control Consortium^19^, and European controls genotyped by the Laboratory of Neurogenetics (LNG) at the National Institute on Aging (NIA), National Institutes of Health (NIH). This data was then meta-analyzed with 1,782 PD+pRBD cases and 131,250 controls from 23andMe Research Institute pRBD differs from iRBD in the fact that the former is diagnosed using the RBD Single-Question Screen (RBD1Q), which has a high sensitivity (93.8%) and specificity (87.2%) for detecting RBD in PD^20^, while in iRBD the diagnosis is confirmed by overnight polysomnography Both study cohorts from the previous RBD GWAS have been described in detail.^17^ The second cohort consists of an additional 721 iRBD patients and 761 controls, again including individuals of European ancestry recruited by the IRBDSG. Demographics for the cohort can be found in Supplementary Table 1. European ancestry was confirmed through principal component analysis (PCA). We conducted two complementary analyses. Our main analysis was a meta-analysis combining the previous GWAS summary statistics, including iRBD and 23andMe PD+pRBD patients, with the summary statistics from our new cohorts. This analysis is undoubtedly biased towards PD due to the inclusion of PD+pRBD data, and we cannot be confident that RBD onset occurred before or after PD diagnosis in these patients. Thus, we also performed an iRBD-only meta-analysis, excluding the PD+pRBD patients and controls from 23andMe. This approach allowed us to identify genetic associations more specific to iRBD, albeit with reduced statistical power due to a smaller sample size. We will refer to the first analysis as “all-RBD” and the second as “iRBD-only” for the remainder of the manuscript. Informed consent forms were administered and signed by all participants before entering the study, and the study protocol was approved by the appropriate institutional review boards.

### Genome-wide association study

For the new samples that were not included in the previous GWAS, genotyping was performed on the OmniExpress GWAS array for 446 cases and 380 controls and on the Neurobooster array for 275 cases and 381 controls, according to manufacturer’s protocols (Illumina Inc). Quality control was performed with plink v1.9 as previously described (https://github.com/neurogenetics/GWAS-pipeline)^21^. Briefly, samples were removed for excess heterozygosity (inclusion criteria of -0.15 <= F <= 0.15), call rate (missingness > 95%), genetic vs. reported sex mismatches, relatedness closer than 3^rd^ degree relatives (pihat > 0.125), and non-European ancestry based on HapMap3 PCA. Variants were excluded for variant missingness (missingness > 95%), notable differences in missingness between cases and controls (p < 1e-04), haplotype missingness (p < 1e-04), and deviation from Hardy-Weinberg equilibrium in controls (p < 1e-04). Imputation was performed on cleaned data with the TOPMed Imputation Server using the TOPMed reference panel r3 and default settings.^22^ Logistic regression of RBD disease status was performed in plink v1.9 on soft call variants (R^2^ > 0.3) with a minor allele frequency (MAF) greater than 0.01. Covariate adjustments were included for age, sex, and the top 5 principal components to adjust for subpopulation differences among the samples. Variants in *GBA1*, N370S and E326K, that were below our MAF threshold but relevant for the analysis based on previous associations with RBD, were manually added. Quality control, imputation, and regression were performed for the OmniExpress and Neurobooster array data separately. The summary statistics for our new datasets were then meta-analyzed with each other and with the summary statistics from the previous RBD GWAS to increase the statistical power to observe associations of smaller effect sizes, using a fixed-weight meta-analysis in METAL.^23^ Genome-wide significance was defined as a *P* < 5e-08. Secondary independent loci within GWAS hits were determined using GCTA-COJO with default parameters.^24^ Linkage disequilibrium statistics were calculated with LDlink.^25^

### Association testing for PD and DLB risk loci

To determine if any established PD or DLB risk loci were associated with RBD risk, we used the independent GWAS-significant loci from the Nalls *et al*.^26^ PD GWAS and largest DLB GWAS^27^ published to date. Variants that were associated with PD and DLB in these GWASs were extracted from the summary statistics of the all-RBD and iRBD-only meta-analyses, and multiple-testing correction was applied to the p-value threshold for the number of variants present in each disease analysis (PD, Bonferroni-corrected *P* threshold of 0.05/108 = 0.00047; DLB, Bonferroni-corrected *P* threshold of 0.05/5 = 0.01).

### Investigation of new loci in PD and DLB GWASs

To assess if newly associated RBD-risk loci play a role in other *α*-synucleinopathies or measures of PD progression, we examined these new loci in GWASs of PD^26^, DLB^27^, PD with and without RBD^28^, age at onset of PD^29^, and scores for motor, cognitive, and composite progression^30^. In this analysis we included all SNPs within 500 kb upstream and downstream of the lead variant in the RBD-associated locus. Motor progression scores were a combination of random-effect slope values from mixed effect models of MDS-UPDRS III, MDS-UPDRS II, and Hoehn and Yahr stage scores against age at onset, cohort, and gender. Cognitive scores used the same methodology with the Montreal Cognitive Assessment (MoCA), semantic verbal fluency, and MDS-UPDRS item 1.1 scores. All scores from both motor and cognitive analyses were taken together for composite progression. Regression slopes were combined with PCA. More information can be found in the original publication.^30^ Association statistics for *RCOR1* variants from the DLB, PD, and PD with and without RBD GWASs, were visualized with linkage disequilibrium (LD) Manhattan plots created in LocusZoom (http://locuszoom.org/).^31^

### Rare variant burden testing in PD and DLB cohorts

Whole genome sequencing data was obtained for PD and DLB cohorts through the Accelerating Medicines Partnership – Parkinson’s disease (AMP-PD) study group v3 release (https://amp-pd.org/). Initial quality control was performed by AMP-PD as previously described (https://amp-pd.org/whole-genome-data), and additional quality control was applied to both cohorts as described for our genotype data above. Individuals of European ancestry were retained for analyses, determined through PCA. Genes within 500 kb upstream and downstream of the lead variant in new RBD-associated loci were retained for the analysis. Gene regions were extracted using plink v1.9^21^, ± 300 base pairs from the start and end of each gene determined using the UCSC Genome Browser^31^. To test for associations of rare variants (MAF < 0.01), which standard regression analyses are underpowered to detect, we performed optimized sequence Kernel association tests (SKAT-O, R program) in each cohort^32^. This included tests for all rare variants, all missense variants, all likely loss of function variants (splicing, frameshift, and stop-gain), all functional variants (missense and loss of function), and all variants with a CADD phred score greater than 20 indicating the top 1% of predicted deleterious variants. Analyses in both PD and DLB cohorts were adjusted for sex, age, and the top 5 principal components.

### Fine-mapping new loci with SusiE and FINEMAP

We performed fine-mapping analyses to identify the most likely causal variant behind the association of new RBD-risk loci. First, we performed conditional and joint analysis with a p-value threshold of 5e-05 on our iRBD cohort to determine if there were any secondary independent signals^24^. Then we created a LD matrix using the same cohort for all variants within 1 Mb of the lead variant in the locus with plink v1.9^21^. We performed fine-mapping analysis on variants with MAF > 0.01 using both FINEMAP (http://www.christianbenner.com)^33^ and SuSiE (https://github.com/stephenslab/susieR)^34^, with default parameters.

### Colocalization analysis

Colocalization analysis was used to assess whether genetic variants associated with new RBD-risk loci also influence gene expression levels (eQTLs) in brain tissue. We performed coloc (version 4.0.1; https://github.com/chr1swallace/coloc)^35^ using the single causal variant assumption (coloc.abf). Cis-eQTLs were obtained from all GTEx v7 brain tissues (https://gtexportal.org/home/). We extracted all variants 500 kb upstream and downstream of the lead variant in new significant loci. Expression levels of new loci in various tissues were explored with the GTEx browser (https://gtexportal.org/home/).^36^ Colocalization assesses five hypotheses, which have been described previously in detail.^37^ Significant hits in our colocalization analysis are those with a posterior probability of hypothesis 4 (PPH4) greater than 0.8.

## Results

### Genome-wide association study identifies new risk locus

We performed a case-control GWAS in the OmniExpress and NeuroBooster datasets separately, and then meta-analyzed these with the previous RBD GWAS^17^ summary statistics for a total of 3,564 cases and 140,393 controls. We found the genomic inflation factor and QQ plot of the meta-analysis to be acceptable, indicating minimal systematic bias (*lambda* = 1.07, *lambda_1000_* = 1.005, Supplementary Fig. 1).

We found a novel association with the *RCOR1* locus in our main analysis (rs35785423, odds ratio [*OR]* = 1.18, *95% CI* = 1.11-1.25, *P* = 3.79e-08, Fig. 1, Table 1, Supplementary Fig. 2, Supplementary Fig. 3). This signal was observed in the previous RBD GWAS with a p-value just below GWAS-significance (*OR* = 1.18, *95% CI* = 1.11-1.25, *P* = 1.32e-07, Table 1), and the signal was boosted by the addition of our new iRBD datasets in which we see a consistent direction and effect size for the *RCOR1* locus (*OR* = 1.2, *95% CI* = 0.94-1.44, *P* = 0.15). We found the association strengths and sizes of effect to be similar between the current and previous RBD GWASs in all previously reported RBD-risk loci (Fig. 1, Table 1, Supplementary Fig. 3). Specifically, the associations with *SNCA, TMEM175,* and *SCARB2* became stronger while the association with *INPP5F* became weaker. In addition to the primary E326K-driven signal (rs12752133), a secondary signal belonging to N370S (rs76763715) was also observed for *GBA1* (Table 1). The association of *SNCA* in the iRBD-only meta-analysis also increased in statistical significance substantially, while retaining a similar effect size as observed in the previous GWAS (Supplementary Fig. 4, Table 1). Although the lead variants in *SNCA* are different between the all-RBD (rs3756059, *OR* = 1.28, *95% CI* = 1.21-1.35, *P* = 3.87e-19) and iRBD-only meta-analyses (rs1372517, *OR* = 1.5, *95% CI* = 1.37-1.63, *P* = 5.08e-20) they are in strong LD, supporting that they are driven by the same underlying variant (*D’* = 0.99, *R*^2^ = 0.93).

**Figure 1.**
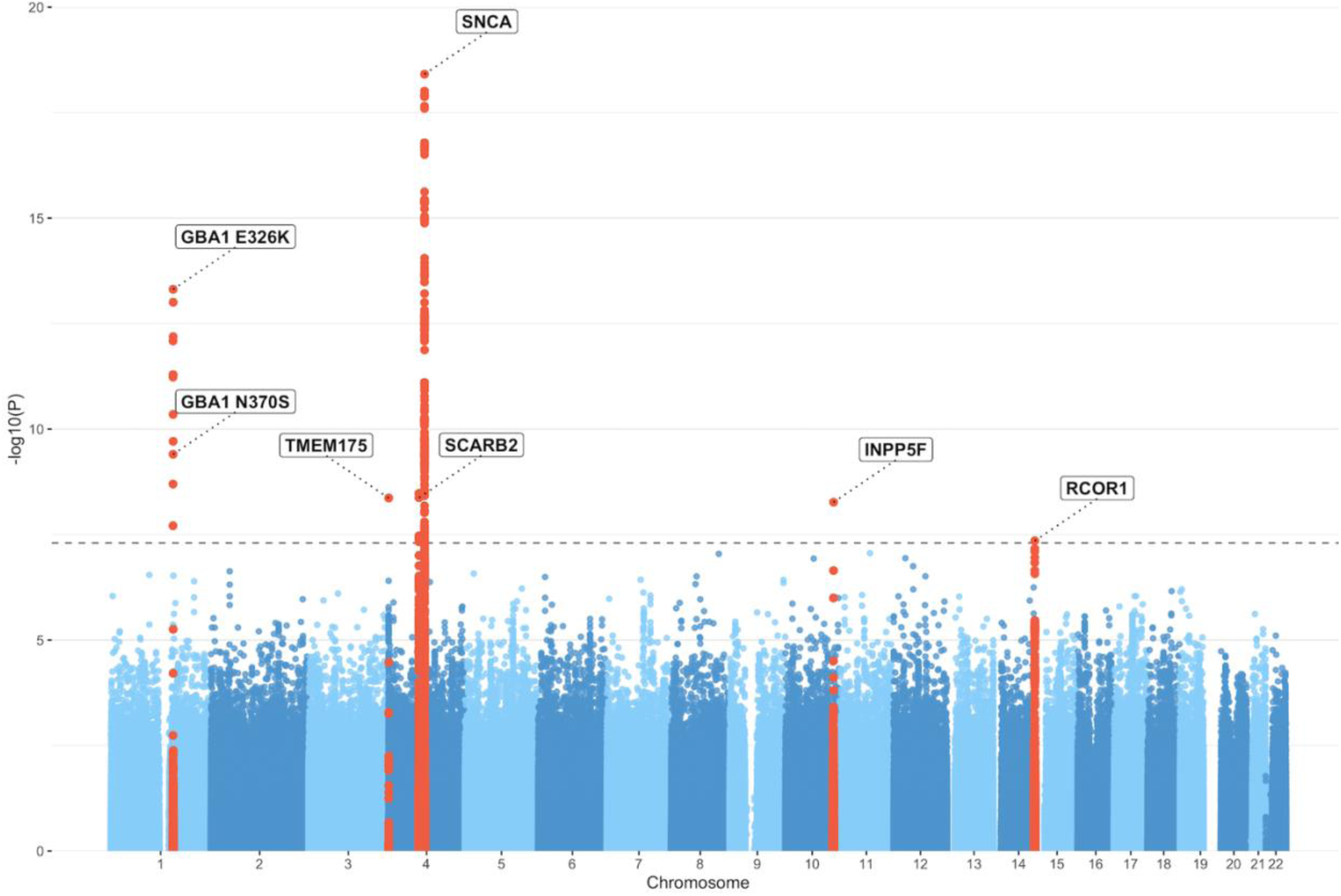
Manhattan plot of log-adjusted p-values at each genomic position. GWAS of RBD disease status, including both iRBD and PD+pRBD patients, adjusted for age, sex, and the top 5 PCs.

**Table 1.** Summary statistics for various RBD-risk GWAS meta-analyses using the new samples cohort.

|  | SNP | Gene | Effect allele | Other allele | New RBD Meta-analysis |  |  |  | Krohn et al. 2022 |  |  |  |
| --- | --- | --- | --- | --- | --- | --- | --- | --- | --- | --- | --- | --- |
| | | | | | MAF | $\beta$ (SE) | OR (95% CI) | P | MAF | $\beta$ (SE) | OR (95% CI) | P |
| all-RBD analysis | rs3756059 | *SNCA* | A | G | 0.5 | 0.24<br>(0.027) | 1.28<br>(1.21-1.35) | 3.87e-19 | 0.5 | 0.23<br>(0.028) | 1.26<br>(1.19-1.33) | 3.02e-16 |
|  | rs12752133 | *GBA1* | T | C | 0.012 | 0.74<br>(0.098) | 2.094 (1.73-2.54) | 4.88e-14 | 0.012 | 0.74<br>(0.098) | 2.094<br>(1.73-2.54) | 4.87e-14 |
|  | rs76763715 | *GBA1* | C | T | 0.004 | 1.067<br>(0.16) | 2.91<br>(2.12-3.99) | 4.5e-11 | 0.003 | 1.045<br>(0.16) | 2.84<br>(2.06-3.92) | 1.68e-10 |
|  | rs34311866 | *TMEM175* | C | T | 0.19 | 0.2<br>(0.033) | 1.22<br>(1.14-1.3) | 4.3e-09 | 0.19 | 0.2<br>(0.035) | 1.21<br>(1.14-1.31) | 4.41e-09 |
|  | rs117896735 | *INPP5F* | A | G | 0.015 | 0.57<br>(0.098) | 1.77<br>(1.46-2.14) | 5.4e-09 | 0.015 | 0.59<br>(0.1) | 1.78<br>(1.48-2.19) | 4.7e-09 |
|  | rs7697073 | *SCARB2* | T | C | 0.34 | 0.17<br>(0.028) | 1.18<br>(1.12-1.25) | 4.23e-09 | 0.34 | 0.16<br>(0.029) | 1.18<br>(1.11-1.25) | 2.21e-08 |
|  | **rs35785423<sup>a</sup>** | ***RCOR1*** | **A** | **G** | **0.29** | **0.16**<br>**(0.03)** | **1.18**<br>**(1.11-1.25)** | **4.45e-08** | **0.29** | **0.16**<br>**(0.031)** | **1.18**<br>**(1.11-1.25)** | **1.32e-07** |
| iRBD-only analysis | rs3756059 | *SNCA* | A | G | 0.5 | 0.4 (0.05) | 1.49<br>(1.37-1.62) | 8.52e-20 | 0.5 | 0.4<br>(0.049) | 1.49<br>(1.36-1.64) | 2.42e-16 |
|  | rs12752133 | *GBA1* | T | C | 0.012 | 0.78<br>(0.18) | 2.18<br>(1.53-3.11) | 1.78e-05 | 0.012 | 0.78<br>(0.18) | 2.18<br>(1.53-3.11) | 1.78e-05 |
|  | rs76763715 | *GBA1* | C | T | 0.005 | 1.26<br>(0.33) | 3.53<br>(1.84-6.77) | 1.48e-04 | 0.003 | 1.18<br>(0.35) | 3.25<br>(1.65-6.41) | 6.79e-04 |
|  | rs34311866 | *TMEM175* | C | T | 0.19 | 0.22<br>(0.054) | 1.24<br>(1.12-1.38) | 5.26e-05 | 0.18 | 0.25<br>(0.06) | 1.28<br>(1.14-1.44) | 3.99e-05 |
|  | rs117896735 | *INPP5F* | A | G | 0.014 | 0.41<br>(0.18) | 1.51<br>(1.056-2.16) | 2.41e02 | 0.014 | 0.45<br>(0.2) | 1.57<br>(1.05-2.34) | 2.68e-02 |
|  | rs7697073 | *SCARB2* | T | C | 0.33 | 0.19<br>(0.045) | 1.2<br>(1.1-1.32) | 3.65e-05 | 0.33 | 0.19<br>(0.05) | 1.2<br>(1.09-1.33) | 2.03e-04 |
|  | **rs35785423<sup>a</sup>** | ***RCOR1*** | **A** | **G** | **0.29** | **0.23**<br>**(0.047)** | **1.27**<br>**(1.15-1.38)** | **1.1e-06** | **0.29** | **0.24**<br>**(0.052)** | **1.28**<br>**(1.15-1.41)** | **2.37e-06** |

RBD, REM-sleep behaviour disorder; GWAS, genome-wide association study; SNP, single nucleotide polymorphism; MAF, minor allele frequency; SE, standard error; OR, odds ratio; CI, confidence interval.
<sup>a</sup>Novel RBD-risk locus, highlighted in bold

### α-Synucleinopathy variants are modifiers of RBD risk

Next, we investigated *α*-synucleinopathy-related variants from the latest PD and DLB GWASs at the time of analysis for associations with RBD risk. We found 8 variants from the PD GWAS^26^ and 3 variants from the DLB GWAS^27^ to be significantly associated with disease status after Bonferroni correction in our all-RBD analysis (Table 2). These include novel RBD associations with PD-risk loci near *STK39* and *SIPA1L2*. The PD-associated variant in the *SCARB2* locus is in moderate LD with the lead *SCARB2* variant in the all-RBD GWAS (*D’* = 0.61, *R*^2^ = 0.29), and was confirmed by GCTA-COJO to be driven by the same signal. Additionally, we identified the PD-associated variant in the *MAPT* locus as having a potentially protective effect for RBD (Table 2). The effect allele for this variant was in LD with a H2 haplotype tagging variant (rs8070723, *D’* = 1.0, *R*^2^ = 0.4), which had a similar significance and effect (*OR* = 0.87, *95% CI* = 0.81-0.92, *P* = 1.48e-05). In the iRBD-only analysis, we observed two PD *SNCA* variants associated with increased iRBD risk (Table 2, Supplementary Fig. 5), which have the opposite direction of effect to that observed in PD, confirming previous observations that in iRBD and PD, *SNCA* variants have opposite effects.^17^ In the analysis of DLB-associated variants (Table 2, Supplementary Fig. 5), we identified the DLB *SNCA* variant to be associated with increased all-RBD and iRBD-only risk. This variant is also in strong LD with the lead variant in our RBD analyses (*D’* = 0.99, *R*^2^ = 0.97) suggesting these encompass the same signal (Supplementary Fig. 6). We also investigated the two Asian-specific PD risk loci identified in the largest Asian GWAS performed to date^38^, finding neither to be associated with RBD in the all-RBD analysis (rs246814, *SV2C*, *OR* = 1.08, *95% CI* = 1.96-2.38, *P* = 0.12; rs9638616, *WBSCR17*, *OR* = 0.98, *95% CI* = 0.93-1.041, *P* = 0.55) or in the iRBD-only analysis (rs246814, *SV2C*, *OR* =1.15, *95% CI* = 0.98-1.34, *P* = 0.084; rs9638616, *WBSCR17*, *OR* = 0.97, *95% CI* = 0.89-1.07, *P* = 0.57). We also investigated the novel PD loci from the multi-ancestry PD GWAS^39^, finding the *IRS2* and *EP300* lead variants to be nominally associated in our all-RBD analysis (Supplementary Table 2). Considering these were not significant in the iRBD-only analysis, this association may be driven by the presence of PD individuals. Neither association remains after applying Bonferroni correction (*P* = 0.0045)

**Table 2.**
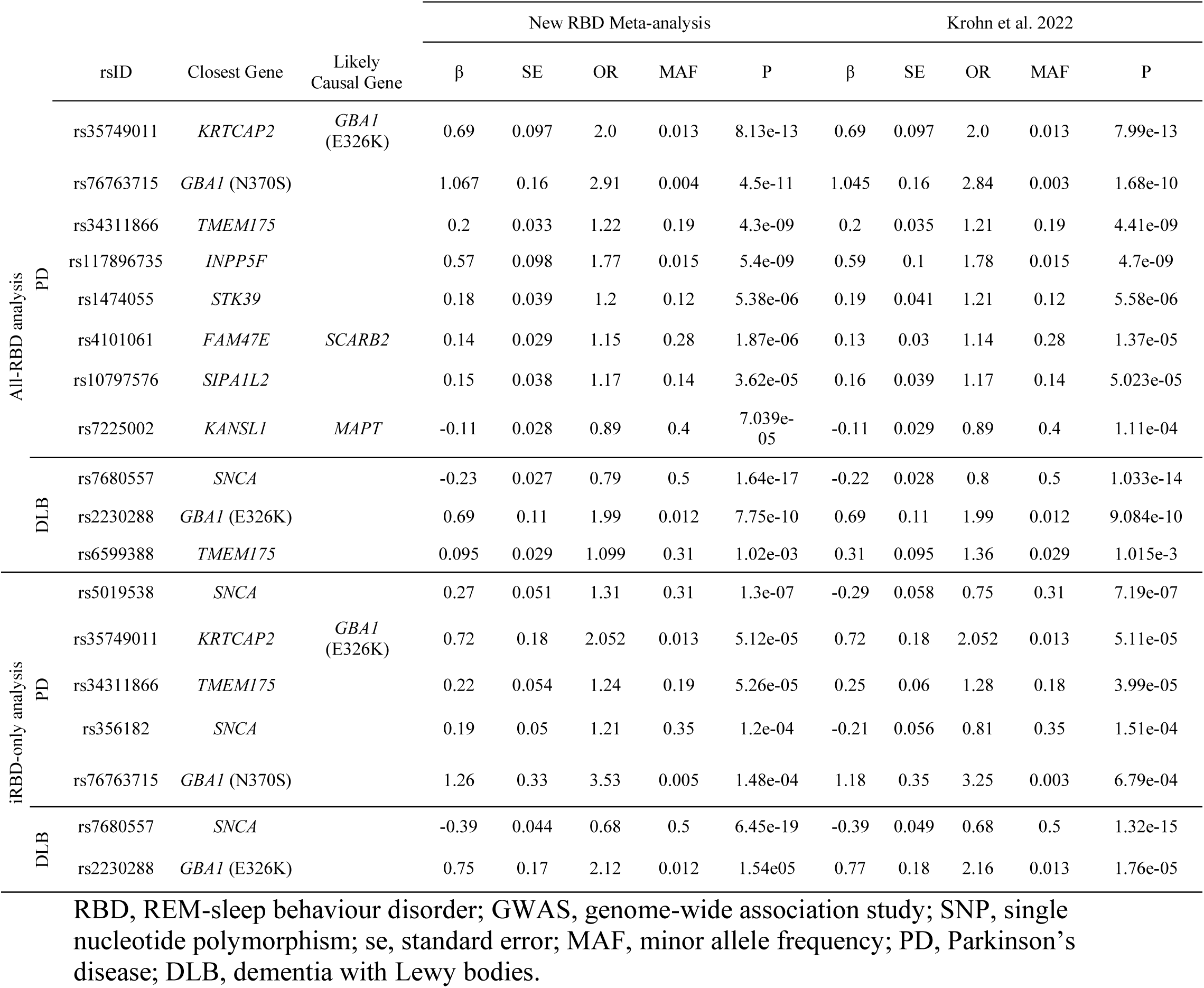
PD- and DLB-risk SNPs associated with RBD risk in main and iRBD-only GWAS meta-analyses.

### Variants in RCOR1 are not associated with other synucleinopathies

We then sought to determine if common (*MAF* > 0.01) and rare variants (*MAF* < 0.01) in the *RCOR1* locus had any evidence for association with disease risk in PD and DLB datasets. In the common variant analysis, all variants within 500kb upstream and downstream of the lead *RCOR1* SNP (rs35785423) were retained for analysis. No variants were associated with PD^26^, DLB^27^, or PD with and without RBD^28^ disease status after Bonferroni correction (Fig. 2, Supplementary Table 3, Supplementary Table 4). Additionally, no variants were associated with PD age at onset^29^, motor progression, cognitive progression, or composite progression^30^ (Supplementary Table 4). To investigate whether rare variants in genes in the *RCOR1* locus were associated with PD or DLB, we performed SKAT-O analyses of the 13 genes within the same ± 500 kb region in PD and DLB whole-genome sequencing datasets from AMP-PD. No rare variant categories in any genes were associated with disease risk in either cohort (Table 3).

**Figure 2.**
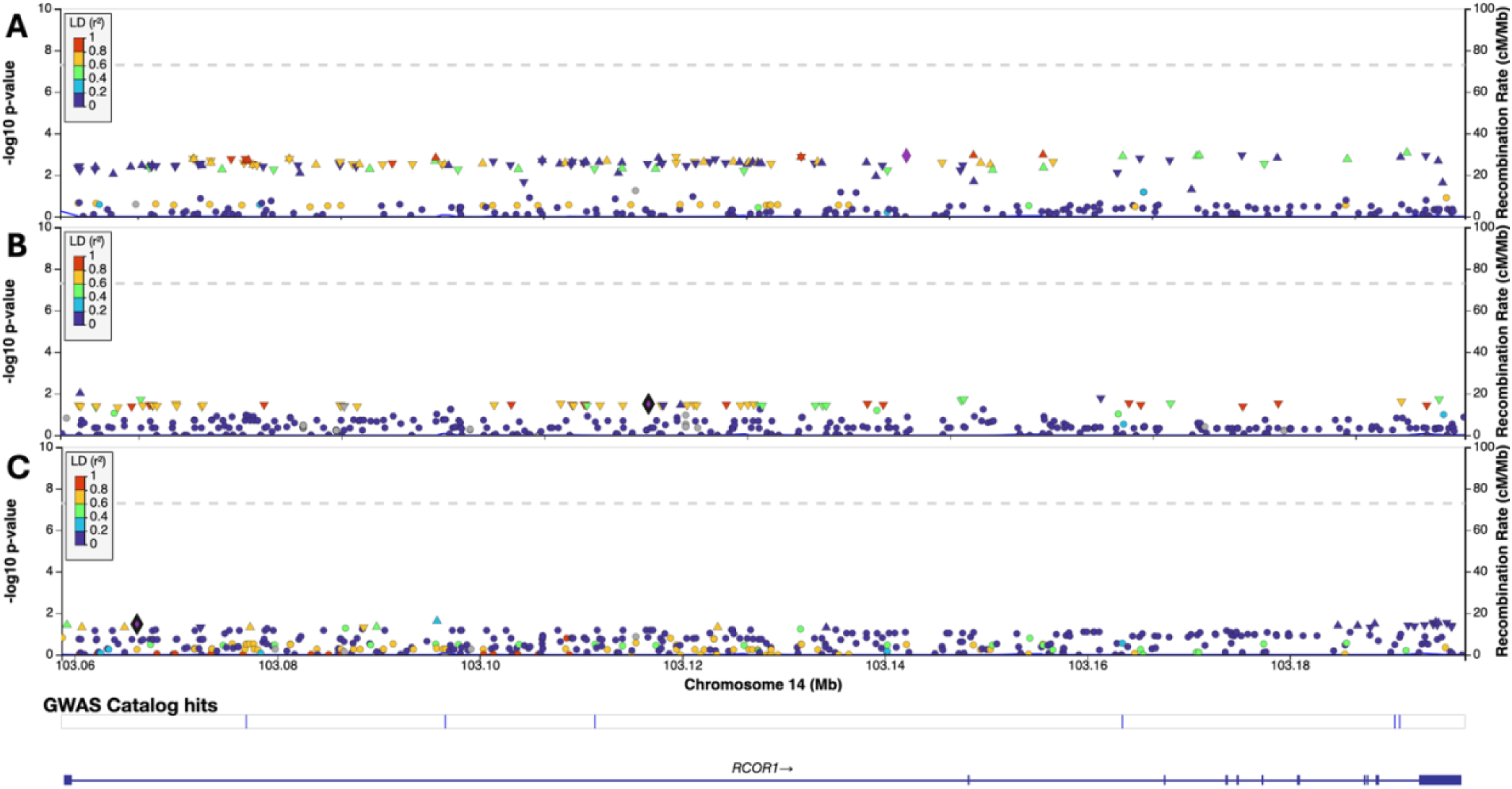
Locuszoom Manhattan plots of log-adjusted p-values at each genomic position for the *RCOR1* gene region. (**A**) GWAS of PD. (**B**) GWAS of DLB. (**C**) GWAS of PD with and without RBD.

**Table 3.**
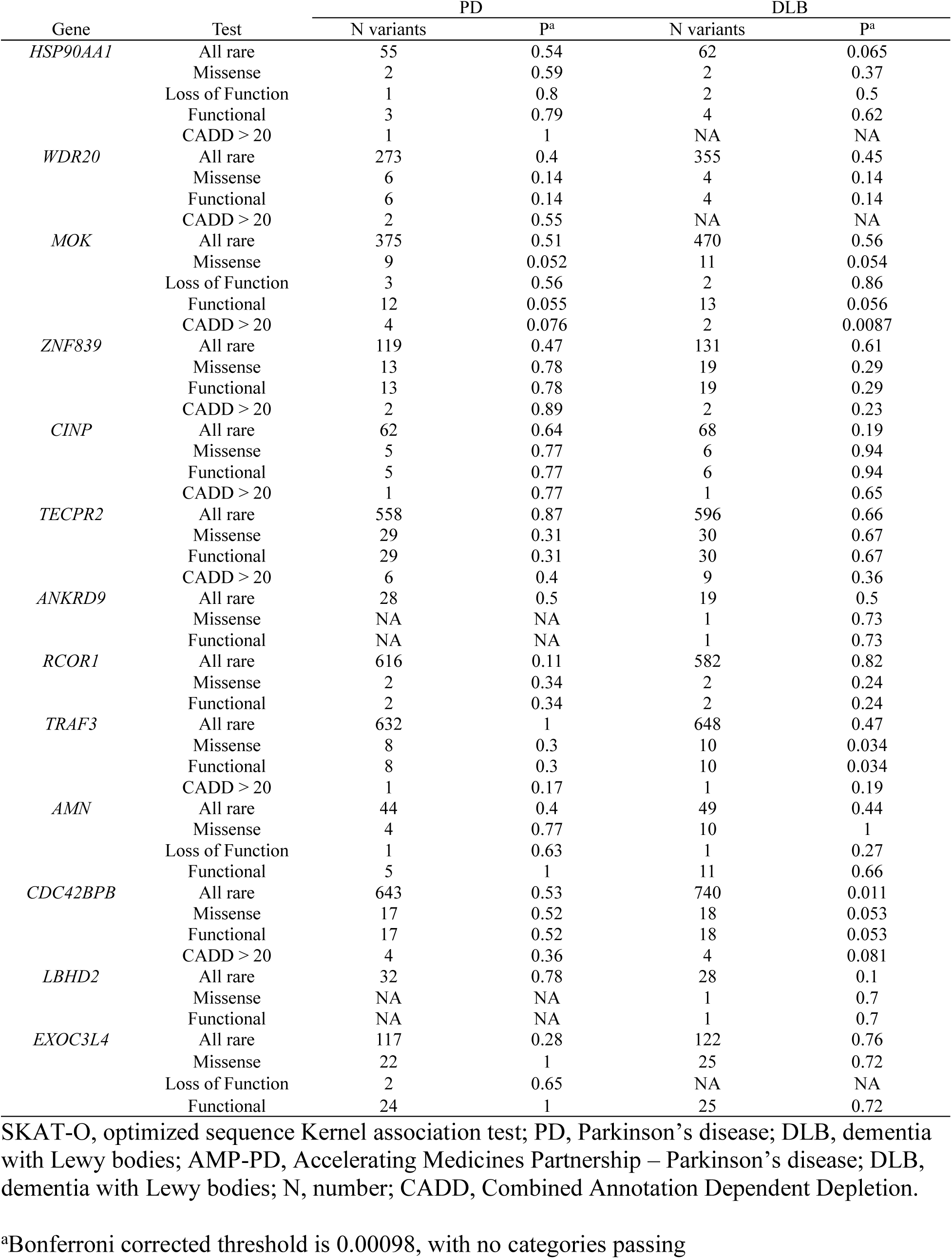
SKAT-O results for the *RCOR1* locus in PD and DLB cohorts from AMP-PD.

### Variants in *RCOR1* drive the association with RBD but are not associated with expression in the whole brain

We performed fine-mapping of the *RCOR1* locus to pinpoint the variants that are likely driving the association with RBD. With conditional and joint analysis, we determined that rs35785423 in intron 2 of *RCOR1* is the only independent signal in the locus. Using FINEMAP, this variant was predicted to be the most likely causal variant in the region with a posterior probability of 0.27. SuSiE predicted two credible sets, the first being led by the same variant (rs35785423, *probability* = 0.33) and the second being led by a variant with a much lower probability (rs7152144, *probability* = 0.093) (Supplementary Fig. 7). The credible sets from both analyses include a high degree of overlap, supporting the variants closely surrounding rs35785423 to be causal for the association in this locus (Fig. 3).

**Figure 3.**
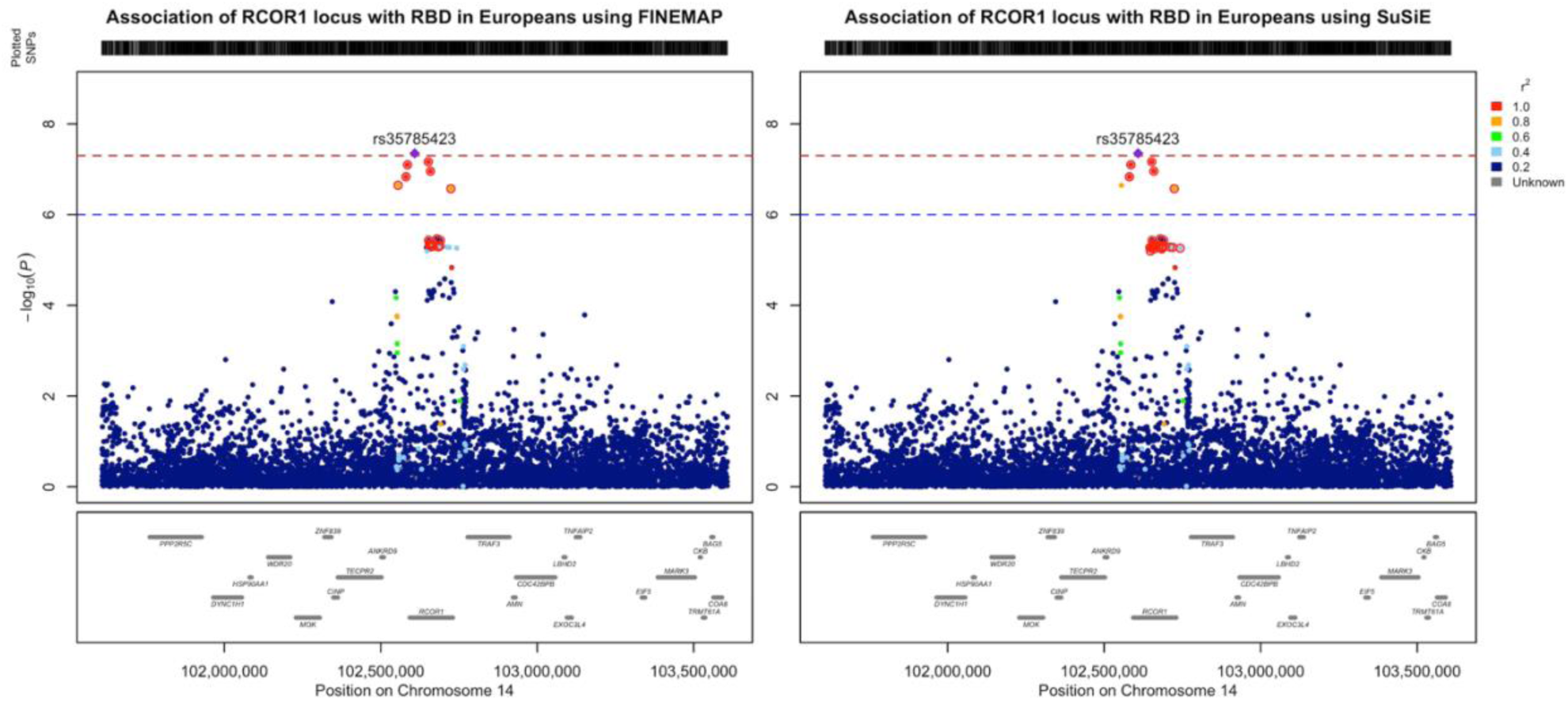
Locuszoom Manhattan plots of log-adjusted p-values at each genomic position for *RCOR1* and the surrounding 1Mb region. Sets of 95% credible SNPs from fine-mapping analysis highlighted in red. (**A**) Results using FINEMAP. (**B**) Results using SuSiE.

Next, we applied colocalization analysis to determine if SNPs in this locus were also associated with tissue-specific brain gene expression using all brain tissue from the GTEx v7 cis-eQTL dataset (https://gtexportal.org/home/). We did not find any evidence for colocalization (*PPH4* > 0.8) with RBD-risk variants in *RCOR1*, or any other surrounding gene, across any of the tissues analyzed (Supplementary Table 5). Notably, we also observed very low expression of *RCOR1* in brain tissue, excluding some expression in cerebellar tissue, potentially explaining the lack of eQTL associations in the area (Supplementary Fig. 8).

## Discussion

The present GWAS identified a new risk locus for RBD in the *RCOR1* region and increased confidence in previously reported associations. Our study also identifies novel PD- and DLB-related loci that potentially affect the risk of RBD, including *MAPT*, *STK39* and *SIPA1L2*.

Although our analyses cannot conclusively identify a causal gene at the novel locus, fine-mapping of the associated region suggests the intron 2 variant within *RCOR1* as the most likely candidate, albeit with low confidence. No evidence was found for comparable signals in nearby genes. Additionally, while no colocalization with brain eQTLs could be detected, and *RCOR1* demonstrates low expression in the majority of brain tissues, its known biological roles make it a plausible candidate to investigate further. *RCOR1* encodes REST Corepressor 1 (RCOR1/coREST1), a component of the coREST complex that regulates gene expression by interacting with REST (RE1-silencing transcription factor) or through REST-independent mechanisms.^40^ This complex has been implicated in the repression of neuronal genes in non-neural tissues, chromatin modification, and neuronal and oligodendrocyte differentiation, with, coREST1 plays an important stage-dependent role in neurogenesis.^40–42^ Thus, alterations to coREST1 could result in the dysregulation of many downstream genes important for proper brain development. Given that coREST1 plays a significant role during neurodevelopment and expression decreases substantially with brain maturation^43,44^, our lack of evidence for a brain eQTL for this locus may be the result of our adult-brain dataset not being optimally suited to capture developmental expression effects. This may also be the result of a lack of power in our dataset, or the intronic variant impacts the disease phenotype through alternative mechanisms not assessed in the present study. It is also notable that we do not observe associations between this locus and PD or DLB, despite these being major outcomes for individuals with iRBD. However, there are several potential explanations for this. Firstly, *RCOR1* could be associated with a non-synucleinopathy aspect of RBD, like sleep processes. Although, this seems unlikely given the role coREST1 plays in neuronal differentiation and neurogenesis.^41,43,44^ We may also fail to see a signal in the PD, DLB and PD with and without RBD GWAS analyses due to the signal being diluted from the presence of PD and DLB patients that did not have prodromal iRBD. Future research focusing on functional aspects of this locus will be required to determine if and how it contributes to the development of RBD.

In addition to the novel association at the *RCOR1* locus, we also observed a reduction in the strength of the association at *INPP5F* following the inclusion of our new iRBD datasets. As a well-established PD risk locus, the decreased significance of *INPP5F* following an increase in the proportion of iRBD patients suggests that its signal in the all-RBD meta-analysis may be primarily driven by the PD with pRBD subgroup, rather than representing a true iRBD risk locus. This will need to be confirmed with additional studies in larger iRBD cohorts, when they become available.

Our study revealed additional associations with known PD and DLB risk loci. We detected several *SNCA* signals in our all-RBD and iRBD-only meta-analyses, which have consistent sizes and directions of effect to what was observed in the previous RBD GWAS and appear to be driven by the same underlying signal. The DLB *SNCA* variant was strongly associated in the all-RBD and iRBD-only analyses and is driven by the same signal as the lead *SNCA* variants in both. The PD-loci in *SNCA* were associated with increased disease risk in the iRBD-only analysis, and displayed an opposite direction of effect to what has been reported in PD. Notably, these associations were no longer significant after adding the PD with pRBD patients in the all-RBD analyses, likely due to the opposite directions of effect in PD and iRBD creating enough phenotypic heterogeneity that it dilutes these SNCA signals. While the PD *SNCA* variant (rs356182) reported in the previous GWAS^17^ appears to be independent of the DLB- and RBD-associated *SNCA* signal identified in our present analyses, the additional PD *SNCA* SNP (rs5019538) displays moderate correlation with the DLB- and RBD-associated *SNCA* locus and may represent a common *SNCA* signal between PD, DLB and RBD. We identified an association with a variant near *FAM47E*, which is in strong LD with the lead RBD-associated *SCARB2* variant. Conditional analysis confirms that these are driven by the same signal in the *SCARB2* locus. Two additional RBD-risk loci were identified from PD variants, including variants near *STK39* and in *SIPA1L2*. *STK39*, or serine threonine kinase 39, plays a role in neuroinflammation and intestinal inflammation and is associated with PD risk in European and Han Chinese populations.^26,45,46^ Similarly, the *SIPA1L2* (signal-induced proliferation-associated 1 like 2) locus is also associated with PD in European and Han Chinese populations^26,47^, but little is known of the mechanism underlying this.^48^ Although these genes appear to be the most likely cause for the SNP associations based on proximity, further research of different genes in these loci and their functional impact in neurodegeneration is needed to identify the genes driving the associations in these loci.

Perhaps the most interesting new RBD locus from the analysis of known PD^26^ and potential DLB^49^ locus is the *MAPT* variant associated with decreased risk, with the observed size and direction of effect being relatively consistent with what has been reported for PD.^26^ Previous research on *MAPT* in RBD yielded mixed support for the locus, with a Spanish cohort finding an association of the protective PD-related variant while a European cohort did not.^16,50^ However, a recent study reported that levels of plasma pTau181, the protein encoded by *MAPT*, are predictive for iRBD phenoconversion to DLB.^51^ Notably, the *MAPT* variant identified in our analysis tags the H2 haplotype, which is known to be associated with decreased PD risk.^52,53^ In contrast, the H1 haplotype is associated with increased risk and potentially cognitive decline in PD, although with less confidence.^53^ This finding of a protective association with the H2 haplotype consistent with previous reports in PD, as well as new reports of pTau181 predicting RBD phenoconversion, provide additional support for the involvement of *MAPT* in RBD and suggest it may play a more significant role than previously thought. Our findings emphasize the need for thorough investigations of *MAPT* at both the genetic and functional level, using larger cohorts with more diverse ancestry, to help unravel the role *MAPT* might play in RBD.

The present study has several limitations. First, we only included individuals with European ancestry. Performing these analyses in diverse populations could uncover more of the genetic basis of RBD and potentially identify ancestry-specific associations. However, unfortunately we do not presently have a large enough cohort in other ancestries to perform a well-powered analysis. Next, we were not able to analyze the specific disease subgroups of RBD, primarily iRBD patients that convert to PD compared to those who convert to DLB. Although we are collecting data to perform this analysis in the future, at the present moment our cohort does not have enough power to detect any significant associations. This study will be important to perform in the future in order to identify RBD loci that can act as markers to help identify type of conversion before it occurs. Lastly, a large number RBD patients utilized in the previous RBD GWAS^21^ are PD patients with pRBD, meaning our study is biased towards RBD-to-PD converters.

In conclusion, we identified *RCOR1* as a novel, potentially RBD-specific risk locus, providing new insights into the genetic architecture of the disease. Despite finding minimal evidence to uncover the underlying mechanism behind this association, our findings lay the groundwork for future research investigating this gene and its role in RBD. Additionally, we discovered several new RBD risk loci shared with PD and DLB, and notably provide new support for *MAPT* as a protective factor. Replication in larger, more diverse cohorts and functional studies will be essential to confirm these associations and elucidate their biological significance.

## Data availability

Code for analyses used in the present study can be found at https://github.com/gan-orlab/RBD_GWAS_2025/. Code used for the fine-mapping analysis can be found at https://github.com/daria-nikanorova/Fine-mapping_SCARB2_CTSB.^54^ Code used for the colocalization analysis can be found at https://github.com/RHReynolds/RBD-GWAS-analysis/. The summary statistics for the main meta-analysis including 23andMe data can be obtained by qualified researchers from 23andMe upon completion of a data access request (https://research.23andme.com/dataset-access/). The summary statistics for the iRBD-specific analysis are publicly available in the GWAS Catalog (https://www.ebi.ac.uk/gwas/). Details on access to summary statistics from the previous Krohn *et al.*^17^ GWAS can be found in the respective publication. The brain eQTL datasets used for colocalization analysis were obtained from the GTEx Portal on 04/28/2025 and can be downloaded from https://gtexportal.org/home/.

## Acknowledgements

Data used in the preparation of this article were obtained from the Accelerating Medicines Partnership® (AMP®) Parkinson’s Disease (AMP® PD) Knowledge Platform. For up-to-date information on the study, visit https://www.amp-pd.org. ACCELERATING MEDICINES PARTNERSHIP and AMP are registered service marks of the US Department of Health and Human Services. This project was supported by the Global Parkinson’s Genetics Program (GP2). GP2 is funded by the Aligning Science Across Parkinson’s (ASAP) initiative and implemented by The Michael J. Fox Foundation for Parkinson’s Research (https://gp2.org). For a complete list of GP2 members see https://gp2.org. We would like to thank the research participants and scientists at the International RBD Study Group (IRBDSG) for contributing to this study. We would also like to thank the research participants and employees of 23andMe Research Institute for making this work possible. We thank Meron Teferra for her assistance. Z.G.O. is supported by the Fonds de recherche du Québec—Santé (FRQS) Chercheurs-boursiers award and is a William Dawson Scholar. E.N.S is supported by a graduate student award from Parkinson Canada. J.-F.G holds a Canada Research Chair in Cognitive Decline in Pathological Aging.

## Funding

This study was financially supported through grants from the Galen and Hilary Weston Foundation and the Michael J. Fox Foundation (MJFF). Additionally, the G-Can (GBA1-Canada) Initiative, an open-science collaborative initiative aimed at addressing GBA1 mutation-based Parkinson’s disease, has made contributions to this research. G-Can is supported by The Hilary and Galen Weston Foundation, Silverstein Foundation, and J. Sebastian van Berkom and Ghislaine Saucier. Samples from Montreal was made available through grants from the Canadian Institutes of Health Research (CIHR). This work was supported in part by the Intramural Research Program of the National Institutes of Health including: the Center for Alzheimer’s and Related Dementias, within the Intramural Research Program of the National Institute on Aging and the National Institute of Neurological Disorders and Stroke.

## Competing interests

Z.G.O received consultancy fees from Lysosomal Therapeutics Inc. (LTI), Idorsia, Prevail Therapeutics, Inceptions Sciences (now Ventus), Neuron23, Ono Therapeutics, Bial Biotech, Bial, Handl Therapeutics, UCB, Capsida, Denali, Simcere, Takeda Pharmaceuticals, Jazz Pharmaceuticals, EG427, Vanqua Bio, Lighthouse, Deerfield and Guidepoint.

## Notes

### Author Declarations

The Institutional Review Board of McGill University gave ethical approval for this work.

