## Supplementary Data for "Genome-wide association study of REM-sleep behaviour disorder identifies new risk loci"

**Supplementary Figures**

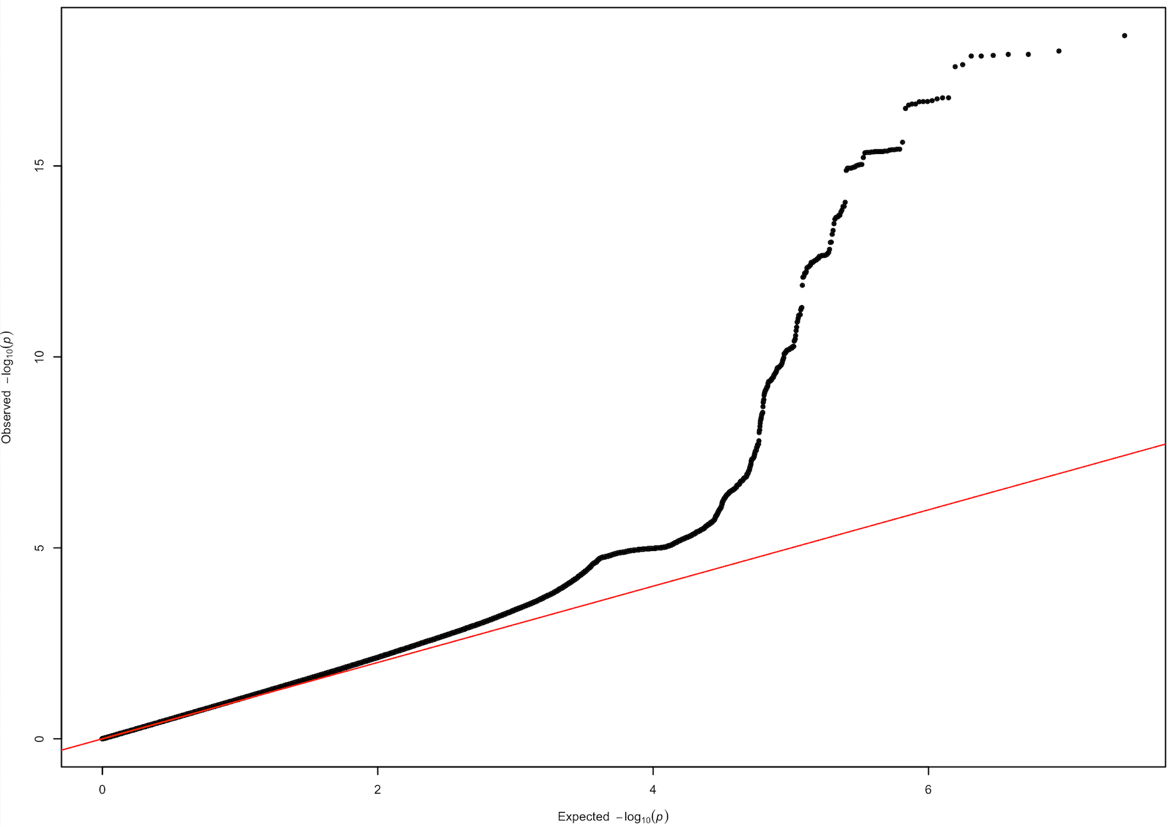

**Supplementary Figure 1.** QQ plot of log-adjusted p-values from the main RBD GWAS meta-analysis, including both iRBD and pRBD patients.

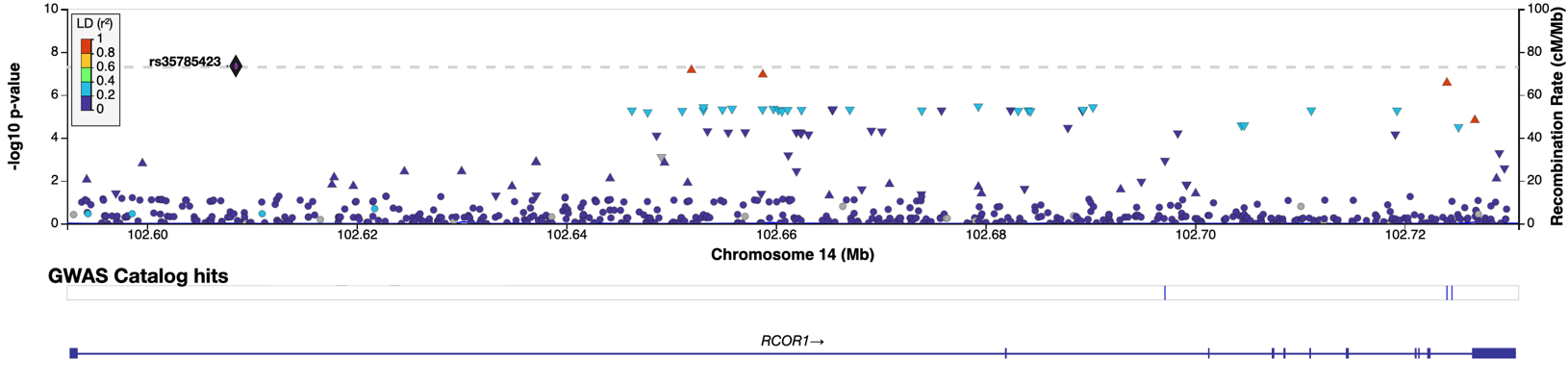

**Supplementary Figure 2.** Manhattan plots of log-adjusted p-values at each genomic position for the *RCOR1* gene region in the main GWAS meta-analysis including both iRBD and pRBD patients.

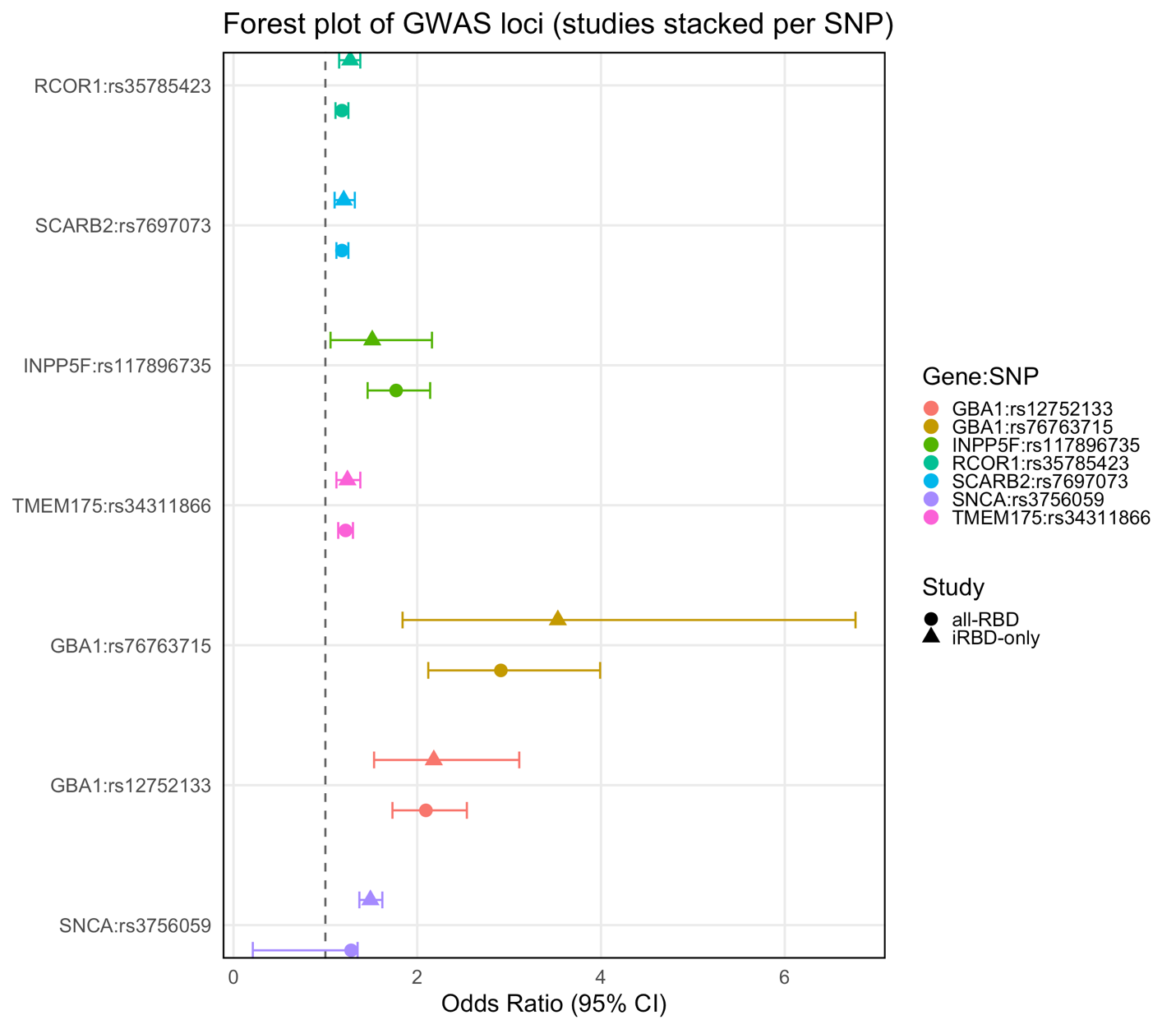
**Supplementary Figure 3.** Forest plot of lead SNPs in GWAS significant loci for both the iRBD-only and all-RBD analyses, representing odds ratios and confidence intervals. SNPs are colour-coded while analyses are shape-coded.

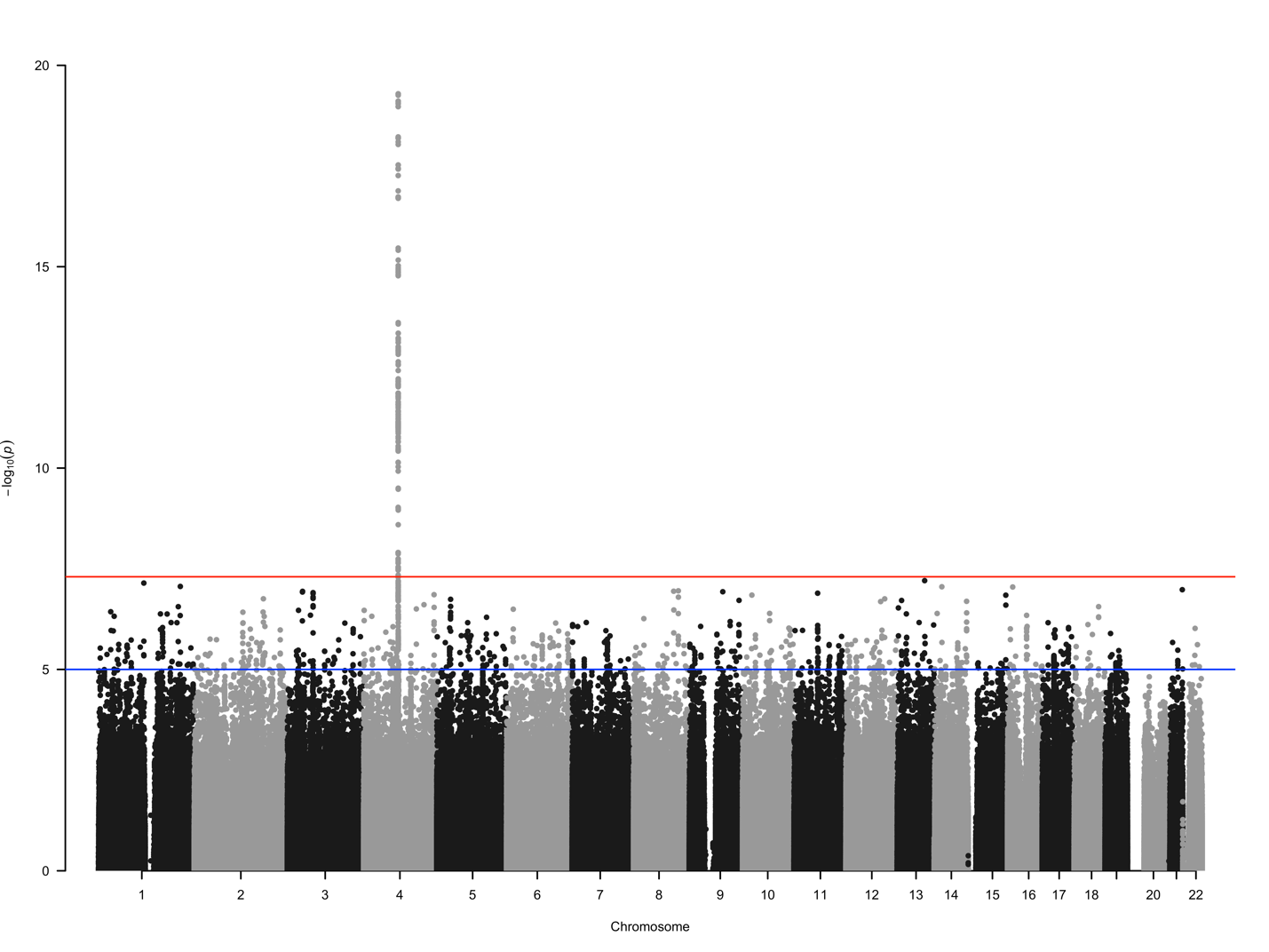

**Supplementary Figure 4.** Manhattan plot of log-adjusted p-values at each genomic position for GWAS meta-analysis using only iRBD patients and controls, adjusted for age, sex, and the top 5 PCs.

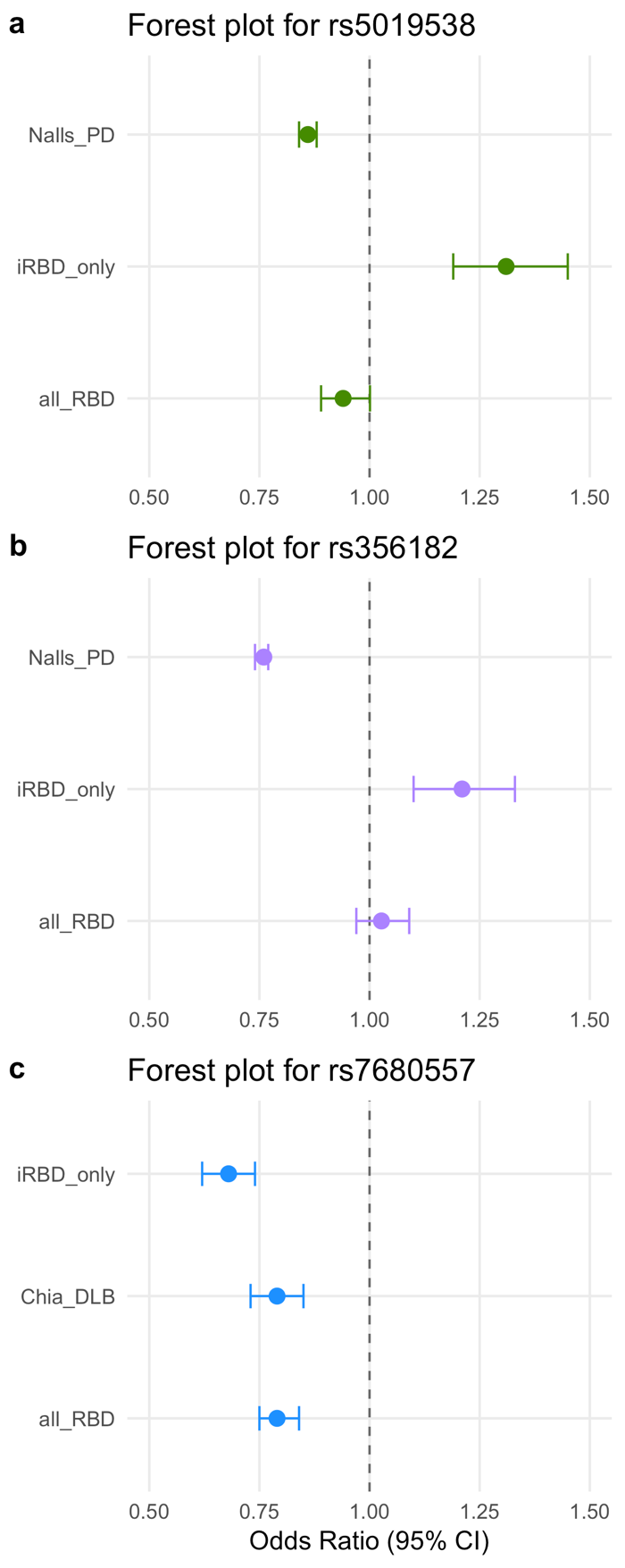

**Supplementary Figure 5**. Forest plot of RBD, PD, and DLB associated lead SNCA variants a) rs5019538, b) rs356182, and c) rs7680557, with points depicting odds ratios and error bars depicting confidence intervals. Points are colour-coded based on the SNP.

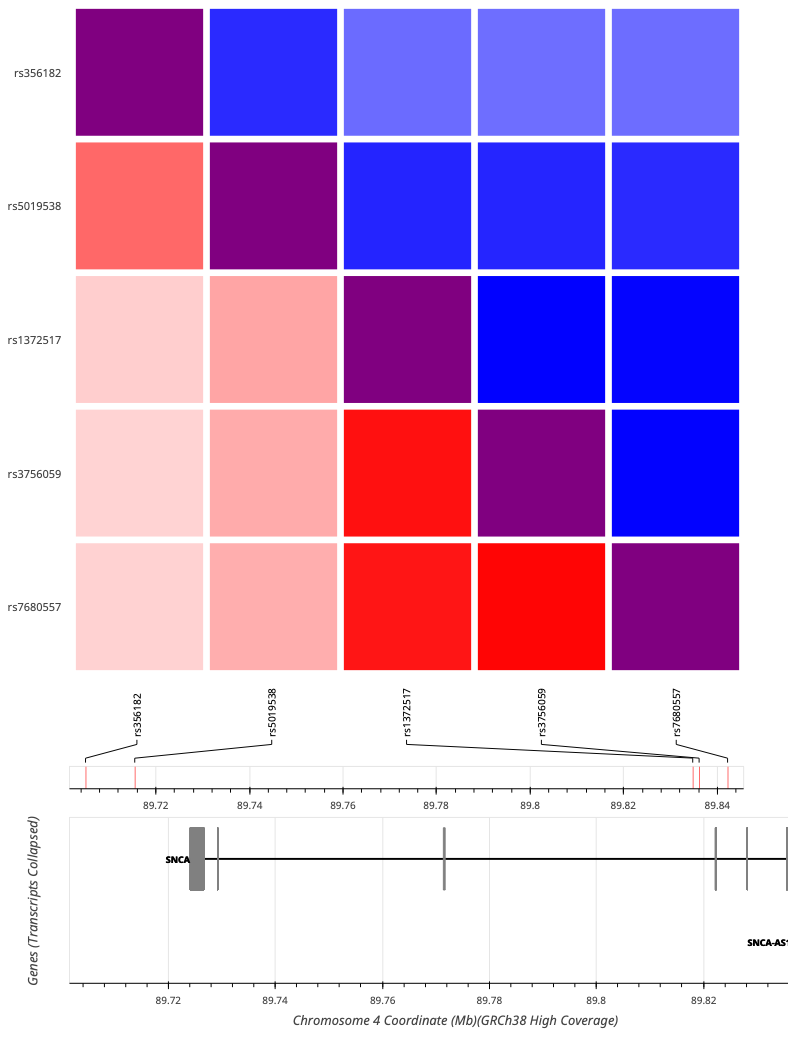

**Supplementary Figure 6.** LDlink linkage disequilibrium heatmap depicting R^2^ in red and D′ in blue for associated *SNCA* SNPs from the main and iRBD-only GWASs.

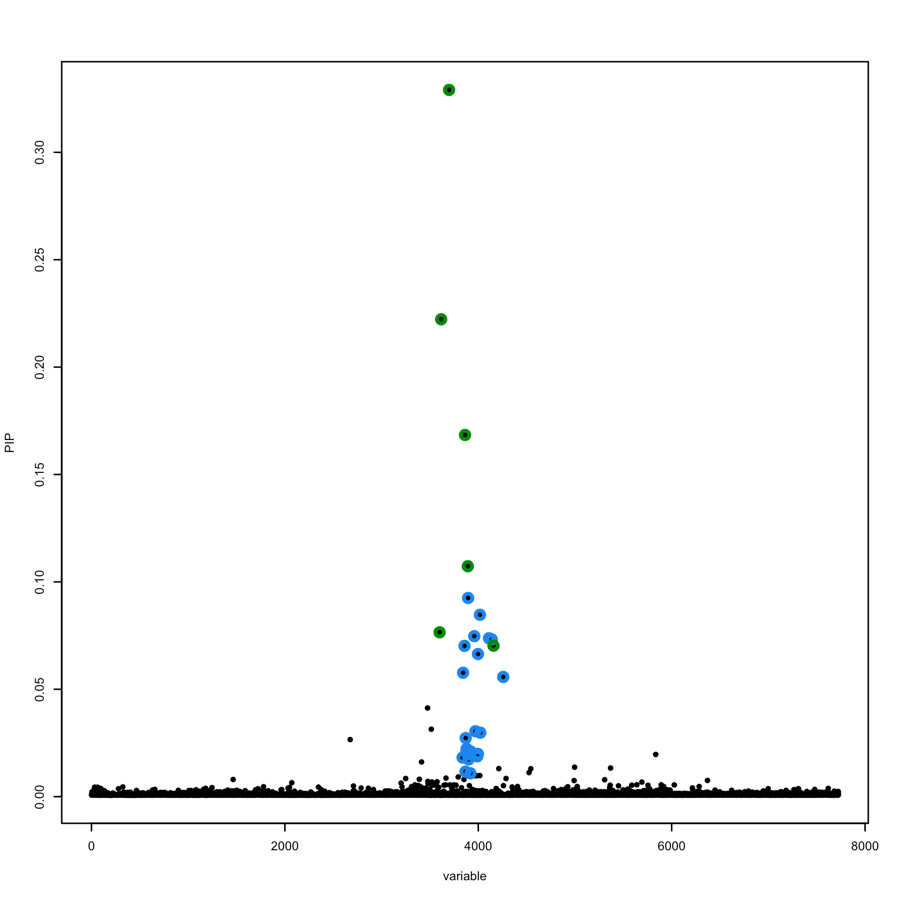

**Supplementary Figure 7.** Manhattan plot of SuSiE PIP values for *RCOR1* and the surrounding 1Mbp region, highlighting the first credible set of causal SNPs in green and the second credible set of causal SNPs in blue.

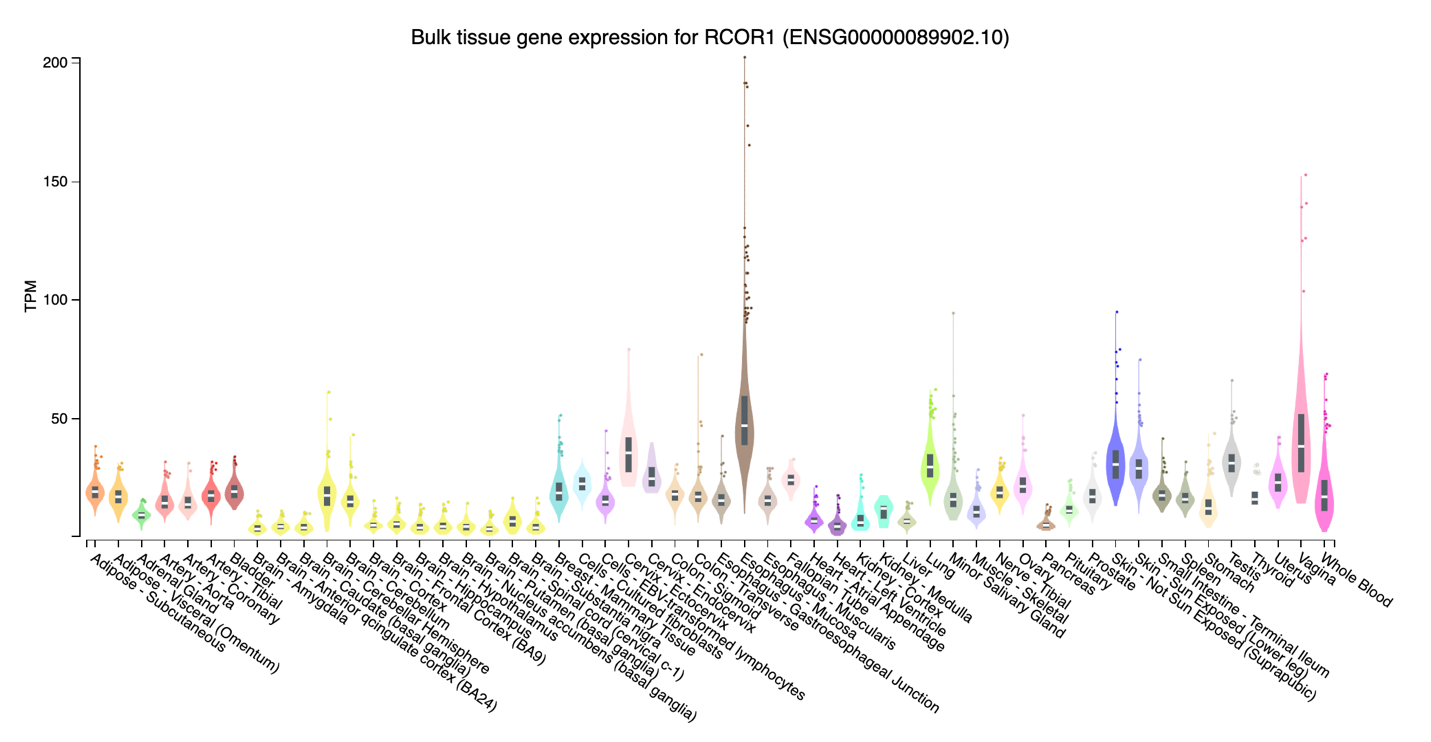

**Supplementary Figure 7.** Gene expression data for RCOR1 in GTEx for each tissue type, with brain tissue being highlighted in yellow.
